# Quantifying lifetime risk for 1,401 infectious diseases across the diabetes spectrum using a Bayesian approach

**DOI:** 10.1101/2025.08.20.25334110

**Authors:** Boomer B Olsen, Martin Tristani-Firouzi, Karen Eilbeck, Mark Yandell, Edgar J Hernandez

**Author notes:** Corresponding author: Boomer Olsen.

## Abstract

Although many diabetes complications have been extensively studied, less is known about the burden of infectious diseases. We developed a Bayesian approach to compare infection risk across 9,476 patients with type 1 diabetes (T1D), 74,270 with type 2 diabetes (T2D), and 32,095 with prediabetes. Patients with T1D, T2D, and prediabetes had multifold increased risk for all organ system- and pathogen-based composite infection outcomes. We also quantified risk for 1,401 individual infection outcomes, finding increased risk for 880 in T1D, 1,047 in T2D, and 991 in prediabetes. Patients had increased risk for well-established diabetes-associated infections (e.g., mucormycosis) and less commonly associated infections (e.g., West Nile Virus encephalitis). Finally, we found disparities in risk across sociodemographic subgroups (i.e., age, sex, ethnicity, ancestry, and insurance status). Our comprehensive findings advance previous research by quantifying risk for wide-ranging infection outcomes across diverse patients with T1D, T2D, and prediabetes through an innovative Bayesian approach.

## 1. Introduction

Diabetes can lead to numerous life-changing complications. Although many diabetes complications have been extensively studied, the burden of infectious diseases across the diabetes spectrum (i.e., type 1 diabetes [T1D], type 2 diabetes [T2D], and prediabetes) is not so clearly defined. Quantifying infection risk is challenging given the breadth and complexity of infectious diseases, especially compared to other diabetes complications. Historically, certain infections have been linked to diabetes via case reports^1^. Examples include but are not limited to mucormycosis^2^, malignant otitis externa^3^, and Fournier gangrene^4^.

Subsequent studies have used more robust methods and electronic health record (EHR) data to characterize features of infection risk. In 2003, a retrospective Canadian study found 2.17-fold skin infection-related hospitalization and 1.92-fold infection-related death among 516,494 people with an unspecified type of diabetes^5^. A prospective Dutch study from 2005 reported 1.24-fold to 1.96-fold risk for several infections (e.g., lower respiratory, urinary tract) among 705 people with T1D and 6,712 people with T2D^6^. In 2015, a retrospective Australian study found 4.42-fold and 1.47-fold infection-related mortality among 85,144 people with T1D and 1,023,838 people with T2D, respectively^7^. In 2017, a retrospective British study reported increased risk for multiple infections (e.g., upper respiratory, pneumonia, urinary tract) among 34,278 people with T2D; a higher most-recent A1c was associated with higher risk^8^. In 2018, another retrospective British study found 2.19-fold risk for 19 infections (e.g., bone, endocarditis) among 5,863 people with T1D compared to 96,630 people with T2D^9^.

Several EHR-based studies have explored whether antidiabetic therapeutics modulate infection risk irrespective of glycemic control. Metformin is a decades-old, widely used antidiabetic therapeutic. A retrospective United States-based study from 2018 reported 0.80-fold pneumonia-related mortality among 7,424 metformin users compared to 7,424 nonusers with T2D^10^. In 2022, a retrospective Taiwanese study found 0.89-fold risk for pneumonia and 0.64-fold risk for respiratory cause of death among 49,012 metformin users with T2D^11^. In 2024, a retrospective Australian study of newer antidiabetic therapeutics reported decreased infection-related hospitalization rates (e.g., pneumonia, urinary tract infections [UTI]), among 99,569 sodium-glucose co-transporter 2 inhibitors users with T2D compared to 186,353 dipeptidyl peptidase-4 inhibitors users with T2D^12^.

Prior studies, including those described here, utilized traditional statistical methods to evaluate connections between infections and diabetes. Bayesian learning and inference are powerful statistical techniques with potential to advance these findings. Bayesian methods enable quantification of the conditional probabilities linking variables (e.g., diabetes and infections) within complex datasets (e.g., EHRs)^13^. We developed a Bayesian approach to query EHR data and compare infection risk across T1D, T2D, and prediabetes. We first computed risk ratios for 1,401 infection outcomes among patients with T1D, T2D, and prediabetes. We then evaluated whether sociodemographic variables modified infection risk. Finally, we explored whether metformin or insulin pump use were associated with changes in infection risk, irrespective of glycemic control.

## 2. Methods

### 2.1. Data source

We used data from the University of Utah Health Enterprise Data Warehouse (EDW)^14^. The EDW contains clinical and sociodemographic information beginning around 1993 for over 1.8 million patients from all university-affiliated healthcare facilities. Data were encoded by ICD-9 and ICD-10 diagnosis codes^15,16^, CPT procedure codes^17^, CUI medication codes (also known as RxNorm)^18^, and LOINC lab test codes^19^. All protected health information data were deidentified. The study was approved by the Institutional Review Board (IRB) at the University of Utah (No. 00138561).

### 2.2. Study design

We explored infection risk utilizing a retrospective cohort design. We included all adults aged 18 years or older in the EDW with a diagnosis of T1D, T2D, or prediabetes. Sociodemographic data are reported in Table 1. We included all infections that occurred after a diagnosis with T1D, T2D, or prediabetes. Risk ratios (RR) for infection outcomes were computed for a cohort of interest (e.g., individuals aged 18 years or older with T1D, T2D, or prediabetes) relative to another cohort of interest (e.g., individuals aged 18 years or older [general population]; individuals aged 18 years or older without T1D, T2D, or prediabetes [nondiabetic cohort]).

**Table 1.** Baseline characteristics for the general population and nondiabetic, type 1 diabetes, type 2 diabetes, and prediabetes cohorts.

|  | **General population**<br>**(N = 1,479,638)** | **Nondiabetic**<br>**(N = 1,373,245)** | **Type 1 diabetes**<br>**(N = 9,476)** | **Type 2 diabetes**<br>**(N = 74,270)** | **Prediabetes**<br>**(N = 32,095)** |
| --- | --- | --- | --- | --- | --- |
| **Sex – N (%)** |  |  |  |  |  |
| Male | 698,571 (47.21) | 646,384 (47.07) | 4,925 (51.98) | 37,754 (50.83) | 13,578 (42.31) |
| Female | 780,632 (52.76) | 726,434 (52.90) | 4,550 (48.02) | 36,509 (49.16) | 18,517 (57.69) |
| **Age – N (%)** |  |  |  |  |  |
| 18-39 | 692,883 (46.83) | 679,273 (49.46) | 3,512 (37.06) | 6,187 (8.33) | 4,855 (15.13) |
| 40-64 | 493,949 (33.38) | 448,148 (32.63) | 3,798 (40.08) | 31,033 (41.78) | 15,348 (47.82) |
| 65-74 | 149,455 (10.10) | 124,044 (9.03) | 1,336 (14.10) | 19,348 (26.05) | 7,075 (22.04) |
| 75-99 | 143,291 (9.68) | 121,719 (8.86) | 833 (8.79) | 17,703 (23.84) | 4,820 (15.02) |
| **Race – N (%)** |  |  |  |  |  |
| Black or African American | 22,573 (1.53) | 19,793 (1.44) | 202 (2.13) | 1,775 (2.39) | 1,101 (3.43) |
| White or Caucasian | 894,968 (60.49) | 819,985 (59.71) | 7,443 (78.55) | 52,550 (70.75) | 21,555 (67.16) |
| Asian | 33,893 (2.29) | 30,111 (2.19) | 165 (1.74) | 2,252 (3.03) | 1,764 (5.50) |
| Native Hawaiian or other Pacific Islander | 14,496 (0.98) | 11,836 (0.86) | 177 (1.87) | 1,992 (2.68) | 722 (2.25) |
| American Indian or Alaska Native | 10,301 (0.70) | 8,369 (0.61) | 146 (1.54) | 1,558 (2.10) | 344 (1.07) |
| Unknown | 503,416 (34.02) | 483,156 (35.18) | 1,346 (14.20) | 14,148 (19.05) | 6,614 (20.61) |
| **Ethnicity – N (%)** |  |  |  |  |  |
| Hispanic | 153,598 (10.38) | 137,981 (10.05) | 992 (10.47) | 10,807 (14.55) | 5,284 (16.46) |
| Not Hispanic | 870,211 (58.81) | 785,915 (57.23) | 7,961 (84.01) | 58,761 (79.12) | 25,088 (78.17) |
| Unknown | 455,832 (30.81) | 449,350 (32.72) | 524 (5.54) | 4,703 (6.33) | 1,724 (5.37) |
| **Insurance – N (%)** |  |  |  |  |  |
| Commercial | 807,973 (54.61) | 759,457 (55.30) | 5,473 (57.76) | 29,667 (39.95) | 18,088 (56.36) |
| Medicaid/Self Pay | 312,547 (21.12) | 296,019 (21.56) | 1,656 (17.48) | 11,383 (15.33) | 4,816 (15.00) |
| Medicare | 219,970 (14.87) | 183,302 (13.35) | 2,028 (21.41) | 29,176 (39.28) | 8,730 (27.20) |

### 2.3. Classification of diabetes

All diagnoses were inferred from ICD codes. The T1D cohort included all individuals billed for T1D (ICD-10 E10). There were 6,165 patients billed for both T1D and T2D (ICD-10 E11); these patients were only included in the T1D cohort. As a result, the T1D cohort represented 0.6% of all patients, which approximates the prevalence of T1D in the United States at 0.5%^20^. Even though T1D and T2D can coexist, double diabetes is infrequently intentionally diagnosed by clinicians^21^. Here, concurrent T1D and T2D diagnoses are more likely due to billing errors that accumulated throughout patients’ exposure to the healthcare system. The T2D cohort includes individuals billed for T2D and excludes those billed for T1D; this cohort was ∼5% of the general population, approximating the expected prevalence of T2D in Utah^22^. The T2D cohort does not exclude those billed for both T2D and prediabetes (ICD-10 R73.03). However, the prediabetes cohort includes individuals billed for prediabetes and excludes those billed for T1D and/or T2D. These inclusion and exclusion criteria ensure each patient is included in a single cohort.

### 2.4. Classification of infection outcomes

We searched the EDW for ICD-9, ICD-10, and LOINC terms indicative of infection diseases, understanding some terms are likely more specific than others. High and abnormal LOINC terms were included, which indicate positive tests for pathogens. In total, we compiled a list of 1,401 terms for analyses (Supplementary Table S1). We then sorted several ICD-9 and ICD-10 terms by organ system, creating nine composite outcomes for cardiovascular, central nervous, gastrointestinal, genitourinary, intra-abdominal, lower respiratory tract, upper respiratory tract, skeletal, and skin and soft tissue infections (Supplementary Table S2). We also sorted LOINC terms by pathogen type, creating composite outcomes for bacterial, fungal, parasitic, and viral infections (Supplementary Table S3). Severity categories were defined by single terms, including bacteremia (ICD-10 R78.81), systemic inflammatory response syndrome (SIRS; ICD-10 R65), severe sepsis (ICD-10 R65.20), and septic shock (ICD-10 R65.21).

### 2.5. Bayesian learning and inference framework

We developed a novel approach to analyze EHR data and quantify infection risk. Our method is rooted in Bayesian learning and inference as described by Barber^23^, enabling dynamic integration of prior clinical knowledge with real-world patient data. The core framework employed conditional dependencies between diabetes status, infection outcomes, and relevant covariates such as sociodemographic variables and clinical comorbidities. We used 100 bootstraps to generate 95% credible intervals (CI) for risk estimates. P-values describe the binomial probabilities that risk for an infection in one subgroup differs from risk in another subgroup, given the data (i.e., the risk ratio). To correct for multiple comparisons, we used the Benjamini-Hochberg adjustment method^24^ to control the False Discovery Rate (FDR), applying a threshold of 0.05. Correlation analyses were performed in GraphPad Prism version 10.0.2 for Mac OS X, GraphPad Software, Boston, Massachusetts USA, www.graphpad.com.

## 3. Results

### 3.1. T1D, T2D, and prediabetes were associated with multifold increased risk for infections of all organ systems by diverse pathogens

The T1D, T2D, and prediabetes cohorts had significantly increased risk for all organ system-based composite infection outcomes compared to the nondiabetic cohort (Fig. 1A). The cardiovascular infection outcome had the greatest risk (i.e., 16.8-fold, T1D; 9.50-fold, T2D; and 3.11-fold, prediabetes). The cardiovascular infection outcome also occurred least often (*n* = 71, T1D; *n* = 319, T2D; and *n* = 31, prediabetes). Composite outcomes with the most events tended to have the lowest risk ratios (e.g., upper and lower respiratory tract infections). Rank-ordered risk, from greatest to least risk, was similar across cohorts (Fig. 1A).

**Fig. 1.**
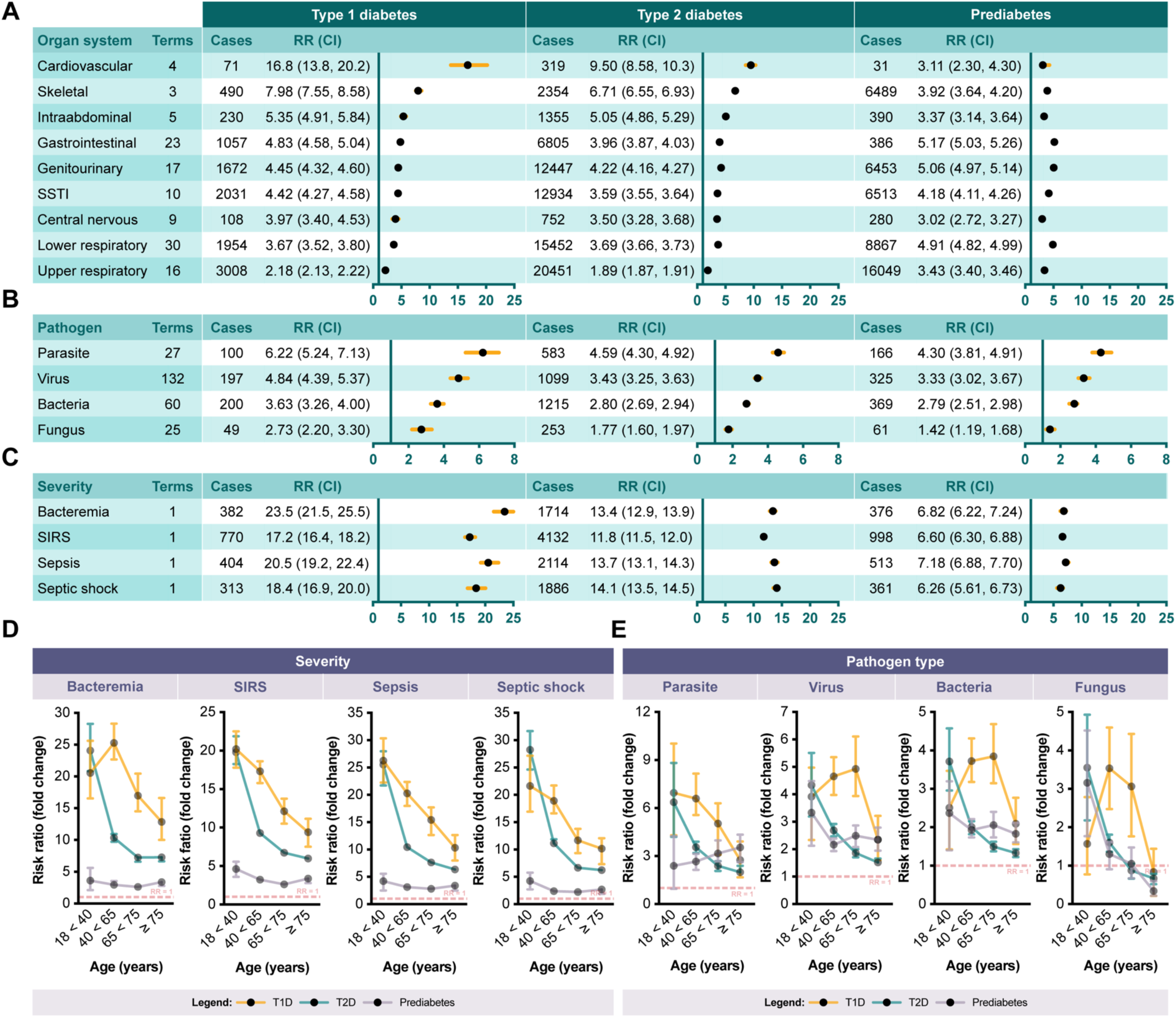
Composite outcome results. A: Forest plots showing risk ratios (RR) and 95% credible intervals (CI) for organ system-based composite outcomes. RR for the type 1 diabetes (T1D), type 2 diabetes (T2D), and prediabetes cohorts are relative to the nondiabetic cohort. B: Forest plots showing RR and 95% CI for pathogen-based composite outcomes compared to the nondiabetic cohort. C: Forest plots showing RR and 95% CI for severe infections compared to the nondiabetic cohort. D: Line plot showing RR with 95% CI for severe infections for T1D, T2D, and prediabetes cohorts compared to age-matched counterparts from the nondiabetic cohort. D: Line plot showing age-stratified RR and 95% for severity-based outcomes for the T1D, T2D, and prediabetes cohorts. RR were computed relative to age-matched nondiabetic counterparts. Terms columns denote the number of terms included in each composite outcome. Cases columns denote the number of cases for each outcome, organized by cohort. E: Line plot showing age-stratified RR and 95% for pathogen-based composite outcomes for the T1D, T2D, and prediabetes cohorts. RR were computed relative to age-matched nondiabetic counterparts. Terms columns denote the number of terms included in each composite outcome. Cases columns denote the number of cases for each outcome, organized by cohort.

The T1D, T2D, and prediabetes cohorts had significantly increased risk for all pathogen-based composite infection outcomes compared to the nondiabetic cohort (Fig. 1B). The parasitic infection outcome had the greatest risk (i.e., 6.22-fold, T1D; 4.59-fold, T2D; and 4.30-fold, prediabetes). The composite outcome with the most events, bacterial infections, had the second lowest risk (i.e., 3.63-fold, T1D; 2.80-fold, T2D; and 3.79-fold, prediabetes). Rank-ordered risk for the pathogen-based composite outcomes was the same across all cohorts (Fig. 1B). Individuals with T1D aged 40 to 75 years tended to have greater risk for parasitic, viral, fungal, and bacterial infections compared to age-matched counterparts with T2D or prediabetes (Fig. 1E).

The T1D, T2D, and prediabetes cohorts had significantly increased risk for bacteremia, SIRS, sepsis, and septic shock compared to the nondiabetic cohort (Fig. 1C). The T1D cohort had the greatest risk for severe infections (i.e., 23.5-fold, bacteremia; 17.2-fold, SIRS; 20.5-fold, sepsis; 18.4-fold, septic shock). When stratified by age, individuals with T1D and T2D aged 18 to 40 years had similar risk for bacteremia, SIRS, sepsis, and septic shock (Fig. 1D). However, individuals with T1D aged 40 years or older had greater risk for severe infections compared to age-matched counterparts with T2D or prediabetes (Fig. 1D). The magnitude of increased risk for the T1D, T2D, and prediabetes cohorts decreased with advancing age (Fig. 1D).

### 3.2. T1D, T2D, and prediabetes were associated with up to 169-fold risk for hundreds of infections

We screened 1,272 ICD, 84 LOINC, and 45 CPT terms related to infection outcomes for increased or decreased risk among T1D, T2D, and prediabetes cohorts compared to the general population (Supplementary Table S1). Out of 1,401 individual terms, we found significantly increased risk for 880 (62.8%) in T1D, 1,047 (74.7%) in T2D, and 991 (70.7%) in prediabetes (Supplementary Table S1). After adjusting p-values to control for the FDR, we still found significantly increased risk for 841 (60.0%) in T1D, 1036 (73.9%) in T2D, and 967 (69.0%) in prediabetes (Supplementary Table S1). Due to low frequencies, there were no changes in risk for CPT terms, which were excluded from further analyses.

Fig. 2 compares fold-change risk for infections across the T1D, T2D, and prediabetes cohorts. Fig. 2A shows a strong positive correlation (*r* = 0.70, *p* < 0.0001) between T1D and T2D, including markedly increased risk for forms of acute and chronic osteomyelitis (ICD-10 M86.17, ICD-10 M86.67). Fig. 2B shows a weaker correlation between T1D and prediabetes (*r* = 0.23, *p* < 0.0001), possibly reflecting distinct comorbidity patterns. Still, T1D and prediabetes shared considerably increased risk for infections such as viral pneumonia (ICD-10 J12.8) and malignant otitis externa (ICD-10 H60.2). Fig. 2C shows a moderate correlation (*r* = 0.34, *p* < 0.0001) between T2D and prediabetes.

**Fig. 2.**
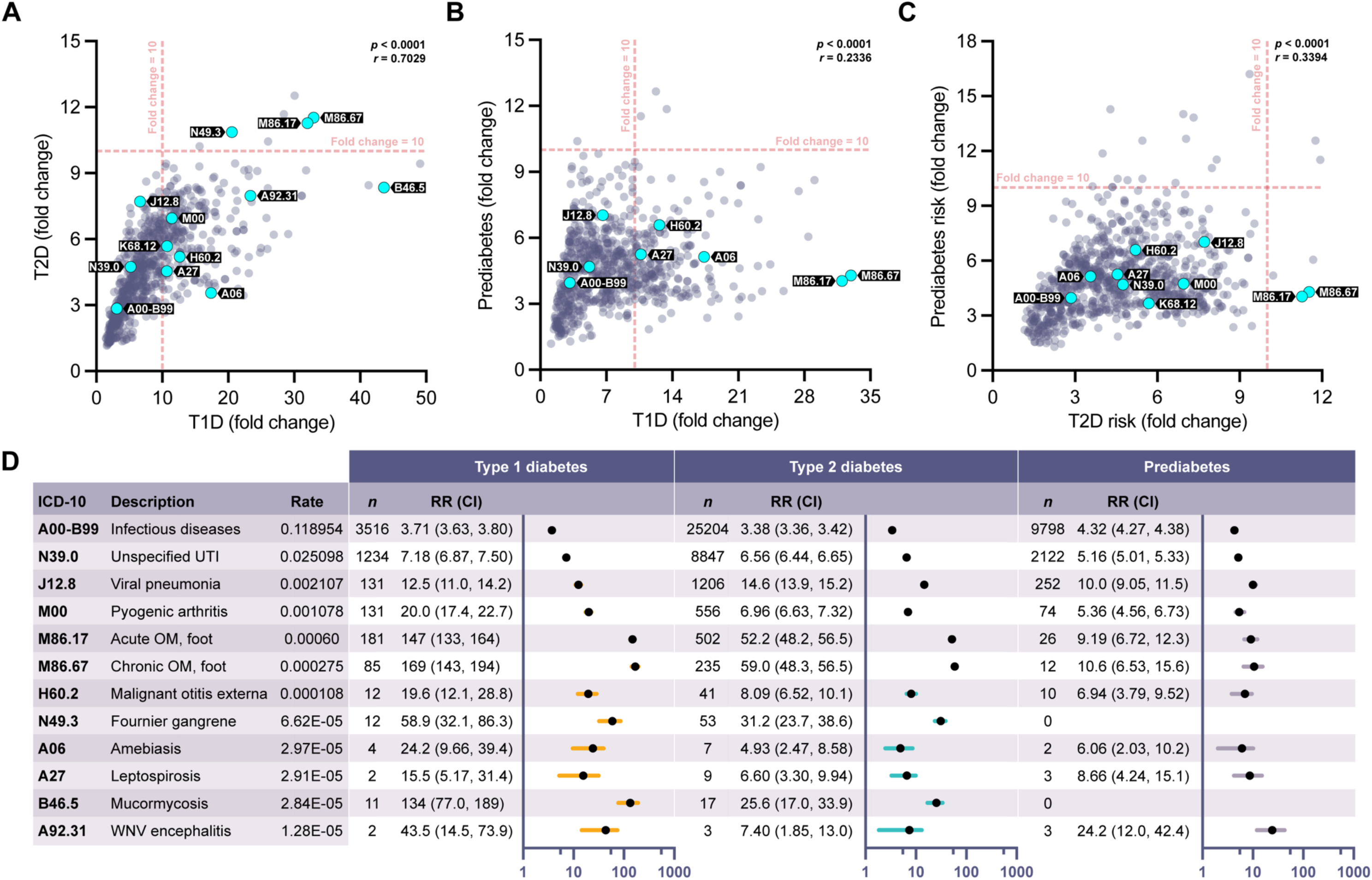
Individual outcome results. A: Scatter plot showing 801 terms with significantly increased risk ratios (RR) shared by the type 1 diabetes (T1D) and type 2 diabetes (T2D) cohorts. RR are relative to the general population. 13 terms of interest are labeled. Values are correlated (*r* = 0.70, *p* < 0.0001). B: Scatter plot showing 771 terms with significantly increased RR shared by the T1D and prediabetes cohorts. RR are relative to the general population. Nine terms of interest are labeled. Values are correlated (*r* = 0.23, *p* < 0.0001). C: Scatter plot showing 883 terms with significantly increased RR shared by T2D and prediabetes cohorts. RR are relative to the general population. 10 terms of interest are labeled. Values are correlated (*r* = 0.34, *p* < 0.0001). D: Forest plots showing RR and 95% credible intervals (CI) for the same terms of interest. RR are relative to the nondiabetic population. Also shown are the rates of each outcome for among the general population and the number of cases (*n*) of each outcome among the cohorts.

The T1D, T2D, and prediabetes cohorts had increased risk for diverse infections compared to the nondiabetic cohort (Fig. 2D). For example, A00-B99 is a composite ICD-10 term that includes multiple "diseases generally recognized as communicable or transmissible" per the Centers for Medicare and Medicaid Services^16^. 176,009 individuals (11.90% of the general population) were diagnosed with A00-B99, making it the most frequent term. The T1D, T2D, and prediabetes cohorts had 3.71-fold, 3.38-fold, and 4.32-fold increased risk for A00-B99, respectively.

As expected, numerous well-established diabetes-related infectious diseases exhibited significantly increased risk ratios across the T1D, T2D, and prediabetes cohorts. Examples include but are not limited to acute osteomyelitis, chronic osteomyelitis, mucormycosis, Fournier gangrene, and malignant otitis externa (Fig. 2D). The T1D, T2D, and prediabetes cohorts had 169-fold, 59.0-fold, and 10.6-fold increased risk for chronic osteomyelitis of the ankle and foot (ICD-10 M86.17), which were the greatest risk ratios for each cohort. T1D, T2D, and prediabetes were also associated with numerous infections that are not historically linked to diabetes. Examples include but are not limited to amebiasis, leptospirosis, and West Nile Virus encephalitis (Fig. 2D), even though these infections occurred infrequently.

### 3.3. Sociodemographic variables and immunosuppression modified risk

We used the most common term (ICD-10 A00-B99) to explore effects of age, sex, insurance status, ethnicity, ancestry, and kidney transplantation and/or immunosuppressive medications. All age groups from the nondiabetic cohort had similar risk for A00-B99 (Fig. 3C). As age increased, T1D subgroups had increasing risk and T2D subgroups had decreasing risk for A00-B99 (Fig. 3C). Individuals with T1D or T2D who identified as female had the highest risk for A00-B99 (Fig. 3D). Among patients with T1D, females had 1.17-fold risk for A00-B99 compared to males (95% CI 1.14-1.21, *p* < 0.0001). Among patients with T2D, females had 1.18-fold risk for A00-B99 compared to males (95% CI 1.16-1.19, *p* < 0.0001).

**Fig. 3.**
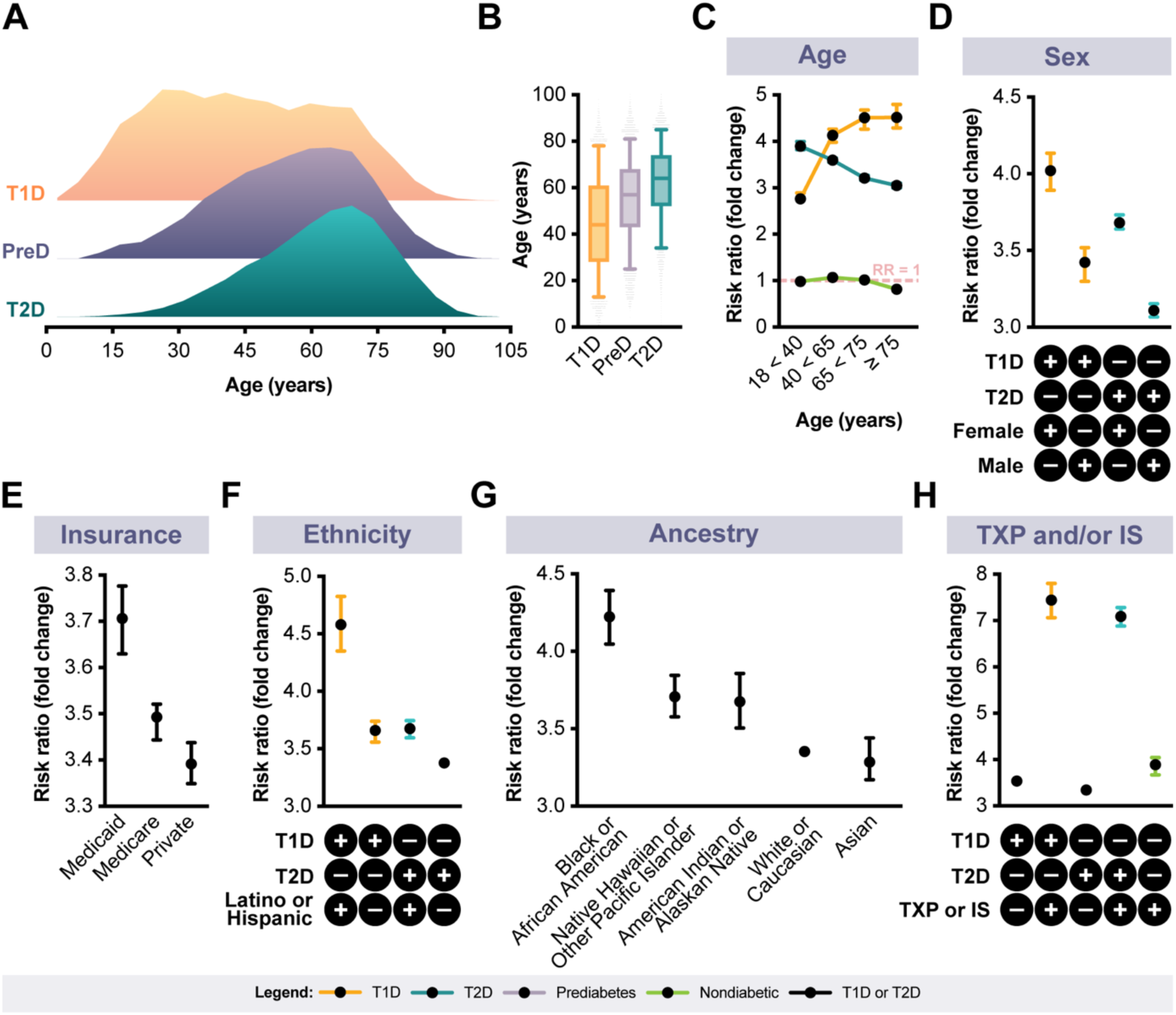
Effects of sociodemographic variables and prior kidney transplantation and/or immunosuppression on risk for ICD-10 A00-B99. A: Ridge plot showing the densities of ages at which type 1 diabetes (T1D), type 2 diabetes (T2D), and prediabetes were first linked to patients. B: Box plot further detailing the ages at which T1D, T2D, and prediabetes were first linked to patients. Boxes denote the median values and interquartile ranges. Whiskers denote 95% credible intervals (CI). C: Line plot showing risk ratios (RR) and CI for A00-B99 across T1D, T2D, prediabetes, and nondiabetic cohorts, grouped by age. RR are relative to the nondiabetic cohort (i.e., all ages). D: Plot comparing RR and 95% CI for A00-B99 among males and females with T1D or T2D. RR are relative to the nondiabetic cohort. E: Plot comparing RR and 95% CI for A00-B99 among individuals with T1D and/or T2D and Medicaid, Medicare, or private insurance. RR are relative to the nondiabetic cohort. F: Plot comparing RR and 95% CI for A00-B99 among individuals with T1D or T2D who do or do not identify as Latino or Hispanic. RR are relative to the nondiabetic cohort. G: Plot comparing RR and 95% CI for A00-B99 among individuals with T1D or T2D and diverse ancestries. RR are relative to the nondiabetic cohort. H: Plot comparing effects of prior kidney transplantation and/or immunosuppressive medications (TXP and/or IS) on RR for A00-B99 among individuals with T1D or T2D. RR are relative to the nondiabetic cohort. Error bars represent 95% CI.

Individuals with T1D or T2D and Medicaid, Medicare, or private insurance had 3.71-fold, 3.49-fold, and 3.39-fold risk for A00-B99, respectively (Fig. 3E). Generally, individuals who identified as Latino or Hispanic had the greatest risk for A00-B99. The Latino or Hispanic subgroup with T1D had the greatest risk, which was 4.58-fold greater than the nondiabetic population (Fig. 3F). Individuals with T1D or T2D who identified as Black or African American, Native Hawaiian or Other Pacific Islander, or American Indian or Alaskan Native had higher risk for A00-B99 than counterparts who identified as White or Caucasian or Asian (Fig. 3G). The Black or African American subgroup had the greatest risk, which was 4.22-fold greater than the nondiabetic population (Fig. 3G).

Prior kidney transplantation and/or use of immunosuppressive medications also modified risk (Fig. 3H). 1,111 individuals with T1D or T2D received a kidney transplant (ICD-10 Z94.0) and/or used tacrolimus (CUI 42316), sirolimus (CUI 35302), and/or mycophenolate (CUI 7145). The presence of kidney transplantation and/or immunosuppressive therapy was associated with 2.14-fold risk for A00-B99 compared to T1D or T2D alone (95% CI 2.08-2.19, *p* < 0.0001). Even in the absence of these terms, individuals with T1D or T2D still had 3.18-fold risk for A00-B99 compared to the nondiabetic population (95% CI 3.16-3.21, *p* < 0.0001).

### 3.4. Change in risk associated with antidiabetic therapeutics

Among 9,476 individuals with T1D, 2,546 had used an insulin pump (ICD-10 Z46.81 and/or Z96.41). We screened 1,356 ICD and LOINC terms to evaluate whether insulin pump use was associated with changes in risk for infection (Fig. 4A; Supplementary Table S4). Both users and nonusers had significantly different risk for 373 terms compared to the general population. Insulin pump users had lower risk than nonusers for 215 of 373 terms (Fig. 4B). Insulin pump users had 0.64-fold risk for infectious arthropathies, 0.65-fold risk for mycoses, 0.73-fold risk for cellulitis and abscesses, 0.81-fold risk for pneumonia and influenza, and 0.92-fold risk for bacterial and viral infections compared to nonusers (Fig. 4C).

**Fig. 4.**
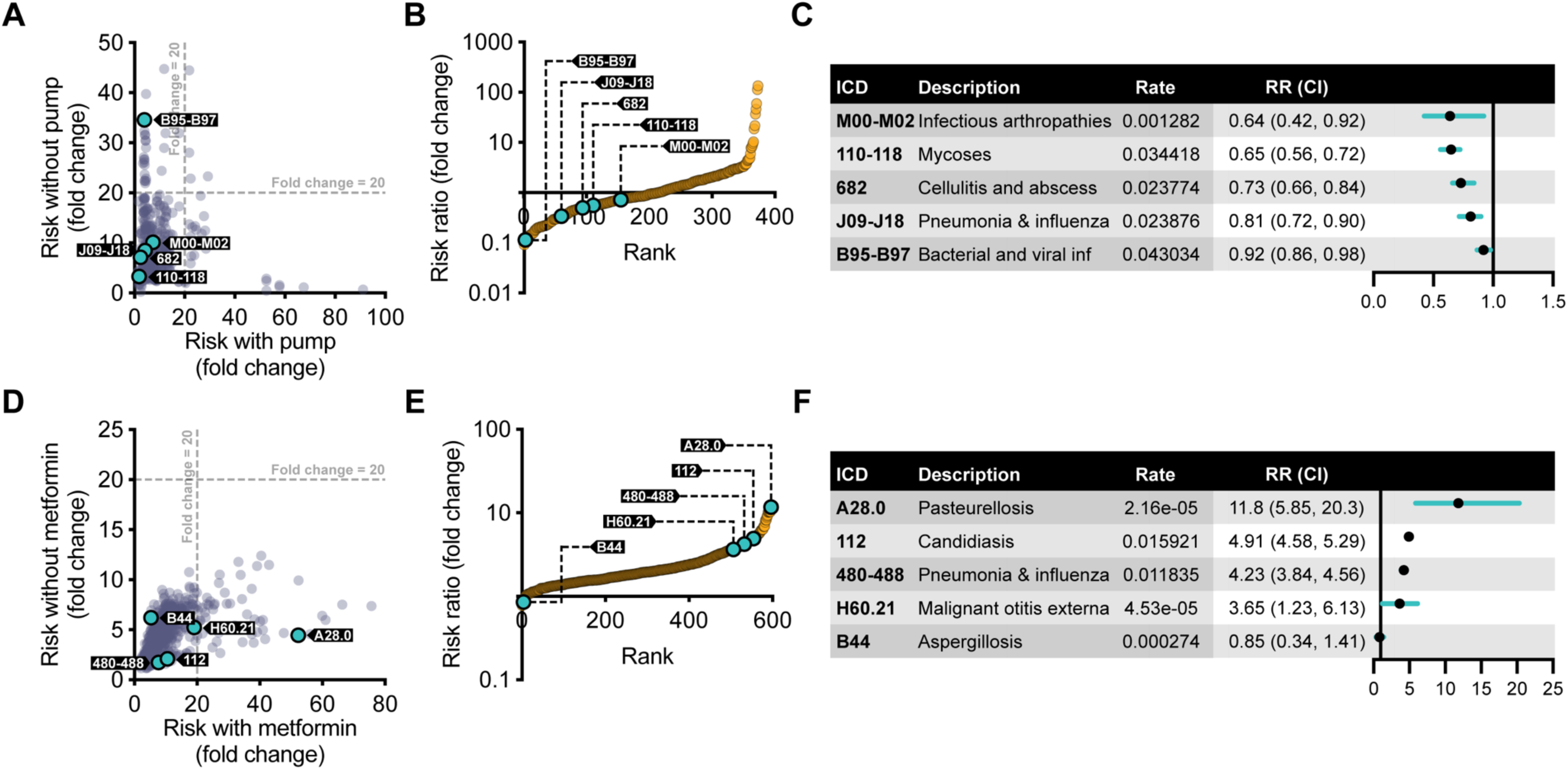
Changes in infection risk associated with insulin pump or metformin use. a, Scatter plot showing significant risk ratios (RR) shared by individuals with type 1 diabetes (T1D) who had or had not used an insulin pump. RR are relative to the general population. Five terms of interest are labeled. B: Plot showing rank-ordered RR. Here, the RR represents RR for insulin pump users relative to RR for counterparts who had not used an insulin pump. C: Forest plot showing RR and 95% credible intervals (CI) for terms of interest among insulin pump users as compared to counterparts who had not used an insulin pump. The rate column denotes the rates of each term among the general population. D: Scatter plot showing significant RR shared by individuals with type 2 diabetes (T2D) who had or had not used metformin. RR are relative to the general population. Five terms of interest are labeled. E: Plot showing rank-ordered RR. Here, the RR represents RR for metformin users relative to RR for counterparts who had not used metformin. F: Forest plot showing RR and 95% CI for terms of interest among metformin users as compared to counterparts who had not used metformin. The rate column denotes the rates of each term among the general population.

Among 74,270 individuals with T2D, 3,425 had used metformin (CUI 6809). We screened the same terms to evaluate whether metformin was associated with changes in risk for infection (Fig. 4D; Supplementary Table S5). Both users and nonusers had significantly different risk for 597 terms compared to the general population. The metformin users had higher risk for 589 of 597 terms compared to nonusers (Fig. 4E). Metformin users had 11.8-fold risk for pasteurellosis, 4.91-fold risk for candidiasis, 4.23-fold risk for pneumonia and influenza, and 3.65-fold risk for malignant otitis externa compared to nonusers (Fig. 4F). Conversely, metformin users had 0.85-fold risk for aspergillosis, which was one of only nine outcomes with decreased risk.

## 4. Discussion

Here, we describe a Bayesian approach used to discover and characterize infectious diseases associated with T1D, T2D, and prediabetes. We first showed that T1D, T2D, and prediabetes are associated with multifold increased risk for composite infection outcomes for each organ system, pathogen, and severity. This advances previous research, which found increased risk for several composite infection outcomes such as lower and upper respiratory tract infections (6), by taking a systematic and comprehensive approach. Interestingly, patterns of ranked risk for composite outcomes were similar across cohorts (Fig. 1A, B, and C), which could reflect biological effects of diabetes on infection risk if not due to confounding variables. Overlapping data cannot explain similarities in risk across cohorts since our exclusion criteria ensure no individuals were included in more than one cohort.

The T1D cohort had greater risk for most composite infection outcomes compared to the T2D and prediabetes cohorts, which is consistent with prior studies comparing only T1D and T2D (9). T1D was generally diagnosed at a younger age (Fig. 3A and B), potentially allowing for more time to accumulate subsequent infections. Compared to age-matched counterparts, the T1D cohort still had greater risk for severe infections (Fig. 1D) and infections by all pathogen types (Fig. 1E). The magnitude of increased risk for T1D, T2D, and prediabetes decreased with older age compared to age-matched nondiabetic counterparts, potentially reflecting that all individuals with advanced age are susceptible for severe infections despite the presence or absence of diabetes.

Risk for the cardiovascular infection outcome was the highest for all three cohorts; it was also the least frequent organ system-based composite outcome. Risk for the upper respiratory infection outcome was the lowest for all three cohorts; it was also the most frequent organ system-based composite outcome. This highlights several challenges in comparing results across composite outcomes. Sampling bias could disproportionally affect infrequent outcomes, such as the cardiovascular infection outcome. Additionally, each unique term included in the composite outcomes likely has variable specificity for a true infection. For example, sinusitis could be used as a diagnosis for both infectious and allergic pathologies, therefore potentially deflating the risk ratio.

The composite outcomes for parasitic, viral, fungal, and bacterial infections included all available positive tests for infections. Thus, these outcomes may be more specific for infections than the organ system-based outcomes, which instead used ICD terms (e.g., sinusitis). Again, the T1D, T2D, and prediabetes cohorts had multifold increased risk for each pathogen-based composite outcome (Fig. 1B). Risk for the parasitic infection composite outcome was significantly higher than risk for the bacterial and fungal infection composite outcomes across all three cohorts. The parasitic infection composite outcome was the least frequent, potentially making it more susceptible to sampling bias. However, these data suggest individuals with T1D, T2D, and prediabetes are particularly susceptible to parasitic infections. Interestingly, a recent meta-analysis also found an association between intestinal parasites and diabetes^25^, even though this is not a well-established relationship.

To circumvent limitations of composite outcomes, we leveraged our robust dataset and powerful computational workflow to screen for associations with individual ICD, LOINC, and CPT terms representing infection outcomes. This is computationally more challenging and, subsequently, has not been performed to the extent done here. We screened 1,401 individual terms for increased risk among T1D, T2D, and prediabetes cohorts compared to the general population (Fig. 2A, B, and C; Supplementary Table S1). We were unable to detect changes in risk for procedures such as incision and drainage due to low CPT event rates. However, the T1D, T2D, and prediabetes cohorts had significantly increased risk for a majority of individual ICD and LOINC terms (Supplementary Table S1). Interestingly, the magnitudes of risk ratios were significantly correlated across all pairwise permutations of the three cohorts (Fig. 2B, C, and D); the strongest correlation was between the T1D and T2D cohorts. This is surprising since prediabetes is a precursor to T2D and is perhaps more closely biologically related. However, we did not account for glycemic control in any analyses, which was likely worse among the T1D and T2D cohorts.

From the screening results, we selected 12 terms of interest for further analyses. We included common (e.g., ICD-10 A00-B99, a predefined composite term including several bacterial and viral infections) and rare terms (e.g., mucormycosis). We included terms that have been historically associated with diabetes (e.g., osteomyelitis) and others that are not obviously linked (e.g., viral pneumonia). For each term, we computed risk ratios compared to the nondiabetic cohort (Fig. 2D). Terms representing acute and chronic osteomyelitis had the highest risk ratios. T1D was generally associated with greater risk than T2D and prediabetes, which is visually represented by Fig. 2A and B. Overall, these data suggest individuals with T1D, T2D, and prediabetes have multifold increased risk for most, if not all, infections -- including common diseases such as viral pneumonia and rare diseases such as West Nile Virus encephalitis.

We then explored effects of sociodemographic variables (i.e., age, sex, insurance, ethnicity, and ancestry) and immunosuppression using the most frequent ICD-10 term (A00-B99). To our knowledge, sociodemographic variables have not been widely integrated into previous research on infection risk among individuals with diabetes. In general, T1D develops at a younger age. Even though we inferred the time of diagnosis using the first billing event for the corresponding ICD-10 term, T1D was still linked to patients at an earlier age than prediabetes and T2D (Fig. 3A and B). This may explain why, at least in part, T1D was associated with greater risk for subsequent infections. All age groups from the nondiabetic cohort had similar risk for A00-B99 (Fig. 3C). However, there was increasing risk for T1D cohort and decreasing risk for the T2D cohort for each age group, as compared to the overall risk for the nondiabetic cohort (Fig. 3C).

Our findings suggest there are disparities in infectious diseases across subgroups of patients with T1D, T2D, and prediabetes. Among the T1D and T2D cohorts, females had higher risk than males (Fig. 3D), Medicaid- and Medicare-insured patients had higher risk than privately insured patients (Fig. 3E), and Latino or Hispanic-identifying individuals had higher risk than their counterparts (Fig. 3F). Black or African American, Native Hawaiian or Other Pacific Islander, and American or Alaskan Native individuals had higher risk than White or Caucasian and Asian populations (Fig. 3G). In contrast, a recent British study concluded that infection risk is similar across ethnic subgroups of patients with T2D and prediabetes, despite some minor differences^26^. Prior kidney transplantation and/or immunosuppressive medications doubled the risk for ICD-10 A00-B99 but did not explain all findings. Even in the absence of transplantation and/or immunosuppression, individuals with T1D and T2D still had greater than threefold risk for ICD-10 A00-B99 compared to the nondiabetic population (Fig. 3H).

Prior studies reported that certain antidiabetic therapeutics, including metformin, are associated with decreased risk for infections such as pneumonia^10,11^. We aimed to evaluate whether metformin was associated with decreased risk for infectious diseases within the EDW. Unexpectedly, metformin users with T2D had higher risk for most infections compared to nonusers, including a term representing pneumonia and influenza (ICD-10 J09-J18; Fig. 4D, E, and F). A significant limitation is that just 4.6% of patients with T2D in the EDW were linked to metformin. We would expect this to be closer to 34.0-59.5% according to a recent survey of metformin use between 1999-2018^27^.

Finally, we were motivated to explore whether insulin pump use was associated with changes in infection risk, particularly skin and soft tissue infections^28^. Insulin pump users with T1D had lower risk for many infections compared to nonusers, including a term representing cellulitis and abscess (ICD-9 682; Fig. 4A, B, and C). Since pumps are expensive, this could be related to sociodemographic variables, among other possible explanations. Importantly, sampling bias could affect data presented in Fig. 4. For example, aspergillosis was an extremely rare event (Fig. 4F), potentially affecting the metformin users by happenstance. Overall, our data were better suited for characterizing risk for infections and effects of sociodemographic variables among the T1D, T2D, and prediabetes cohorts.

## Data Availability

The datasets generated and analyzed during the current study are available in the GitHub repository at https://boomerolsen.github.io. Data are also provided within the manuscript and supplementary information files.

## Code Availability

The underlying code for this study is not publicly available for proprietary reasons.

## Acknowledgements

We thank the members of the University of Utah EDW and Utah Center for High Performance Computing, especially Barry Moore, for computational support and facilitating access to medical records. We also greatly appreciate the valuable discussions shared with Nathan Blue, Scott Watkins, and Marie Couldwell. B.B.O. was supported in part by the Utah Stimulating Access to Research in Residency (StARR) program under NIH Award Number 1R38HL167282-01. M.Y. and M.T.F were supported in part under NIH Award Number U01HL128711. The content is solely the responsibility of the authors and does not necessarily represent the official views of the NIH.

## Author Contributions

B.B.O. and E.J.H. conceptualized the study. B.B.O. performed the data analyses with feedback from all authors, especially E.J.H. and M.Y. M.Y. wrote the code. B.B.O. wrote the paper, which was reviewed and edited by all authors. All authors approved the final version of the manuscript. B.B.O. is the guarantor of this work and, as such, had full access to all the data in the study and takes responsibility for the integrity of the data and the accuracy of the data analysis.

## Competing Interests

The authors have no competing interests to declare.

## Ethics Approval and Consent to Participate

The study protocol was approved by the Institutional Review Board (IRB) at the University of Utah (No. 00138561). The IRB waived the requirement for obtaining informed consent due to the retrospective nature of the study. All methods were performed in accordance with the Declaration of Helsinki.

## Notes

### Competing Interest Statement

The authors have declared no competing interest.

### Author Declarations

IRB of the University of Utah waived ethical approval for this work.

## References

1. Joshi, N., Caputo, G.M., Weitekamp, M.R., Karchmer, A.W. Infections in patients with diabetes mellitus. N Engl J Med. 341,1906–1912 (1999).

2. Smith, H.W., Kirchner, J.A. Cerebral mucormycosis; a report of three cases. AMA Arch Otolaryngol. 68, 715–726 (1958).

3. Zaky, D.A., Bentley, D.W., Lowy, K., Betts, R.F., Douglas, R.G. Jr. Malignant external otitis: a severe form of otitis in diabetic patients. Am J Med. 61, 298–302 (1976).

4. Chernyadyev, S.A., et al. Fournier’s Gangrene: Literature Review and Clinical Cases. Urol Int. 101, 91–97 (2018).

5. Shah, B.R., Hux, J.E. Quantifying the risk of infectious diseases for people with diabetes. Diabetes Care. 26, 510–513 (2003).

6. Muller, L.M., et al. Increased risk of common infections in patients with type 1 and type 2 diabetes mellitus. Clin Infect Dis. 41, 281–288 (2005).

7. Magliano, D.J., et al. Excess Risk of Dying From Infectious Causes in Those With Type 1 and Type 2 Diabetes. Diabetes Care. 38, 1274–1280 (2015).

8. Hine, J.L., et al. Association between glycaemic control and common infections in people with Type 2 diabetes: a cohort study. Diabet Med. 34, 551–557 (2017).

9. Carey, I.M., et al. Risk of Infection in Type 1 and Type 2 Diabetes Compared With the General Population: A Matched Cohort Study. Diabetes Care. 41, 513–521 (2018).

10. Mortensen, E., Anzueto, A. Association of metformin and mortality for patients with diabetes who are hospitalized with pneumonia (Abstract). Eur Respir J. 52, PA2639 (2018).

11. Yen, F.S., Wei, J.C., Shih, Y.H., Hsu, C.C., Hwu, C.M. Metformin use and the risk of bacterial pneumonia in patients with type 2 diabetes. Sci Rep. 12, 3270 (2022).

12. Tan, G.S.Q., et al. SGLT-2 Inhibitor Use and Cause-Specific Hospitalization Rates: An Outcome-Wide Study to Identify Novel Associations of SGLT-2 Inhibitors. Clin Pharmacol Ther. 115, 1304–1315 (2024).

13. Airoldi, E.M. Getting started in probabilistic graphical models. PLoS Comput Biol. 3, e252 (2007).

14. University of Utah ITS Enterprise Data Warehouse. Accessed 26 January 2025. Available from https://risr.hci.utah.edu/helpdocs/ccr/edw/itsedw.html.

15. Centers for Disease Control and Prevention. International Classification of Diseases, Ninth Revision, Clinical Modification (ICD-9-CM). Accessed 26 January 2025. Available from https://archive.cdc.gov/#/details?url=https://www.cdc.gov/nchs/icd/icd9cm.htm.

16. Centers for Disease Control and Prevention. ICD-10-CM Tabular List of Diseases and Injuries. Accessed 26 January 2025. Available from https://www.cms.gov/medicare/coding/icd10/downloads/6_i10tab2010.pdf.

17. American Academy of Professional Coders. What is CPT? Accessed 26 January 2025. Available from https://www.aapc.com/resources/what-is-cpt.

18. National Library of Medicine. RxNorm: Prescription for Electronic Drug Information Exchange. Accessed 26 January 2025. Available from https://www.nlm.nih.gov/research/umls/rxnorm/RxNorm.pdf.

19. Centers for Medicare & Medicaid Services. Logical Observation Identifier Names and Codes (LOINC). Accessed 26 January 2025. Available from https://mmshub.cms.gov/measure-lifecycle/measure-specification/specify-code/LOINC.

20. Fang, M., Wang, D., Selvin, E. Prevalence of Type 1 Diabetes Among US Children and Adults by Age, Sex, Race, and Ethnicity. JAMA. 331, 1411–1413 (2024).

21. Khawandanah, J. Double or hybrid diabetes: A systematic review on disease prevalence, characteristics and risk factors. Nutr Diabetes. 9, 33 (2019).

22. American Diabetes Association. The Burden of Diabetes in Utah. Accessed 26 January 2025. Available from https://diabetes.org/sites/default/files/2023-09/ADV_2023_State_Fact_sheets_all_rev_Utah.pdf.

23. Barber D. Bayesian Reasoning and Machine Learning (Cambridge Univ. Press, 2012).

24. Benjamini, Y., Hochberg, Y. Controlling the false discovery rate: a practical and powerful approach to multiple testing. J R Stat Soc Series B Stat Methodol. 57, 289–300 (1995).

25. Zibaei, M., et al. Intestinal parasites and diabetes: A systematic review and meta-analysis. New Microbes New Infect. 51, 101065 (2022).

26. Carey, I.M., et al. Evaluating Ethnic Variations in the Risk of Infections in People With Prediabetes and Type 2 Diabetes: A Matched Cohort Study. Diabetes Care. 46, 1209–1217 (2023).

27. Fang, M., Wang, D., Coresh, J., Selvin, E. Trends in Diabetes Treatment and Control in U.S. Adults, 1999–2018. N Engl J Med. 384, 2219-2228 (2021).

28. Patel, B., Priefer, R. Infections associated with diabetic-care devices. Diabetes Metab Syndr. 15, 519–524 (2021).

